# Predictors of Interruptions in Antiretroviral Therapy among People Living with HIV in Nigeria: A Retrospective Cohort Study Using the Nigeria National Data Repository

**DOI:** 10.1101/2024.03.08.24303976

**Authors:** Sunday Ikpe, Aliyu Gambo, Rebecca Nowak, John Sorkin, Manhattan Charurat, Timothy O’Connor, Kristen Stafford

## Abstract

This study aimed to identify predictors of time to first interruption in treatment (IIT) and predictors of ever being interrupted in ART treatment among PLHIV in Nigeria using a national longitudinal dataset that covers all PEPFAR-funded implementing partners to inform national strategies to prevent IIT. This retrospective cohort study used data from Nigeria’s National Data Repository (NDR). The NDR is a de-identified longitudinal database of over 1.9 million PLHIV who received ART in Nigeria beginning in 2004 and is owned by the Federal Ministry of Health (FMoH). The NDR contains patient-level demographics, clinic visits, laboratory, and ART prescription and refill data uploaded at least monthly. The data extracted for this study were obtained from electronic medical record systems of 2,226 public facilities offering HIV care in the country. In this study, we investigated the predictors of treatment interruption using data from the national HIV treatment program. We identified sets of predictors of first interruption in treatment using the logistic regression and these to be consistent in predicting time to first interruption including sex, non NRTI drug in ART regimen, recorded HIV viral load, recorded CD4 cell count, WHO clinical staging, functional status, last measured weight, highest education attained, occupation, marital status, year enrolled in care, pre and post surge, pre and post-COVID and residing in a state capital, Lagos, FCT (urban) versus other locations (rural). Age grouping was the only variable that was predictive only for time to first interruption but not for having a first interruption. To reduce the risk of IIT it is important to target interventions preemptively. We have highlighted the need for tailored interventions that address the unique needs of PLHIV in Nigeria. Targeted interventions focusing on those with a combination of risk factors could include education, counseling, supportive services, and monitoring and outreach.

## Introduction

Over 40 years since its emergence, human immunodeficiency virus (HIV) remains a significant public health concern. This is particularly true in sub-Saharan Africa, which bears 67% of the global burden^1–3^, where HIV remains one of the leading causes of death.[1] Antiretroviral therapy (ART) is critical for both individual-level management of HIV infection to preserve or improve health, and to prevent the onward transmission of the virus for epidemic control[2], [3], [4]. While tremendous progress has been made over the last 20 years with the support of the President’s Emergency Plan for AIDS Relief[5] and other international donors to expand access to ART[6], globally 15% of people do not know they are living with HIV, 25% are not on treatment, and 32% are not virally suppressed which inhibits progress towards HIV-1 epidemic control.[1] UNAIDS first announced global targets for progress toward epidemic control in 2014 and then updated them in 2020[7]. The current goal, known as the 95-95-95 target, is for 95% of people living with HIV to be diagnosed, 95% of those who know their status to be on ART, and 95% of those on treatment to be virally suppressed[1]. ART is a lifelong treatment with no cure or vaccine available for HIV.

Based on the 2018 population-based Nigeria AIDS Impact and Indicator Survey, only 47% of people living with HIV have been diagnosed[8]. While the survey revealed that 96% of people who had been diagnosed were on ART, viral suppression among those on treatment was only 81%[9], meaning across the entire cascade only 37% of PLHIV in Nigeria were virally suppressed. Multiple factors may inhibit the achievement of viral suppression targets, as has been observed in Nigeria. These factors may be care or person-related[10]. Interruptions in treatment (IIT) pose a threat to achieving the expected gains of ART for PLHIV and their communities.

Prior studies have been carried out to understand the predictors of IIT and loss to follow-up (LTFU) in not just Nigeria, but in other sub-Saharan countries, including Mali, Ethiopia, and Rwanda, and have identified different sets of predictors including age, baseline CD4 count, sex, marital status, education, access to care, treatment regimens, and treatment fatigue to mention a few[11], [12], [13], [14], [15], [16], [17]. Most of the evidence generated from Nigeria has been based on cohorts from groups of health facilities supported by individual organizations that may employ specific models of care and can also limit geographic representation. These studies have employed only logistic regression or survival analysis as a means of identifying predictors of LTFU or IIT. While each approach is valid, each measures a different outcome. These differences may account for narrow and contradictory findings related to factors driving IIT and limit the generalizability of these studies to other regions[11], [17], [18].

This study aimed to identify predictors of the time to first IIT and predictors of ever being interrupted in ART treatment among PLHIV in Nigeria using a national longitudinal dataset that covers all PEPFAR-funded implementing partners to inform national strategies to prevent IIT.

## Methods

### Design and data source

This retrospective cohort study used medical records from Nigeria’s National Data Repository (NDR). The NDR is a de-identified longitudinal database of over 1.9 million PLHIV who received ART in Nigeria beginning in 2004 and is owned by the Federal Ministry of Health (FMoH)[16]. The data was obtained on the 1^st^ of November 2021 following a University of Maryland Baltimore (UMB) institutional review board (IRB) determination that the research is nonhuman subject research. At no point did we, the authors, have access to any form of personally identifiable information. The NDR contains patient-level demographics, clinic visits, laboratory, and ART prescription and refill data uploaded at least monthly. The data extracted for this study were obtained from electronic medical record systems of 2,226 public facilities offering HIV care in the country.

### Study population and variable definitions

Data from adults ages ≥18 years with a first ART prescription date from January 1, 2015, to March 31, 2021, were eligible for inclusion in the study population. To be included in the analysis, patients had to have had at least two ART visits.

The outcome variable of interest in this study was the first interruption in treatment (IIT), defined as the first instance of no documented pharmacy refill within 28 days after the expiry of their last ART refill, followed by a return to treatment. The variable was derived from the difference between the last refill visit date and the next scheduled refill date which was then binary categorized. Variables of interest included age categories (18 – 24 years and subsequently in 10-year bins), sex (male, female), year of enrollment in care (2015 - 2021), local government area of care (urban, rural), third drug in ART regimen, employment (employed, unemployed, student or retired), marital status, measured and recorded CD4, measured and recorded HIV viral load in copies per ml, WHO clinical staging (including an undocumented level as a proxy for quality of HIV care), functional status (bedridden, ambulatory, working), and last measured weight measured in kilograms. Additionally, a variable to assess the effect of the COVID pandemic was created using a cut point of March 2020 when there was a global lockdown. Similarly, a variable was created to assess the effect of a CDC, Surge initiative[19] to identify more people who were infected with HIV in Nigeria in 2019.

### Statistical analysis

The variables were analyzed considering the frequencies and proportions for categorical variables and the median and interquartile ranges at each of these levels for continuous variables. Chi-square and t-test p-values were reported. Logistic regression was used to estimate the odds of first treatment interruption. Determining potential predictors was based on p-values less than a threshold determined using the Bonferroni approach of dividing 0.05 by the number of variables in the final model. Odds ratios and 95% confidence intervals were estimated for the final selected predictors, along with p-values for these estimates. The outcome of survival analysis was the time to the first interruption event. All individuals who did not experience interruption were censored at the end of the follow-up period while those who met the definition of IIT were censored after the first event. Patients who did not return to care after the interruption event were assumed to have been lost to follow-up or dead and were excluded, restricting the analysis to IIT and patients who continue in care. A long rank test was used to assess differences in time to IIT generated using Kaplan-Meier plots for each categorical variable. Backward elimination was used to determine the variables to be included in a Cox proportional hazards regression model to produce hazard ratios and associated 95% confidence intervals.

The analysis was performed using R programming language version 4 and the survival package.

## Results

The NDR dataset contained records of 4,805,247 people, 32,758,238 clinic visits, 31,612,050 regimen records, and 24,931,718 laboratory results. After merging the datasets, 924,847 adults from 2,204 facilities were eligible for inclusion in the study. The records show an upward trend in enrollment from 89,535 in 2015 to 264,777 in 2020. The 2019 and 2020 surge years had the highest number of enrollees with 510,117(55.20%). The study sample was predominantly women 606,222(65.50%) and aged 25-34 years (Fig 1). The most commonly prescribed ART regimen included integrase strand transfer inhibitors (INSTI) 434,289(47.00%), followed by non-nucleoside reverse transcriptase inhibitors (NNRTI) 369,891(40.00%), protease inhibitors (PI) 5,624(0.60%) and nucleoside reverse transcriptase inhibitors (NRTI). There were 29,648(12.40%) records of undocumented ART. Less than a quarter of the study subjects (215,389, 23.3%) had a recorded CD4. Similarly, slightly over one-third (317,195, 34.3%) had a recorded HIV viral load.

**Fig 1.**
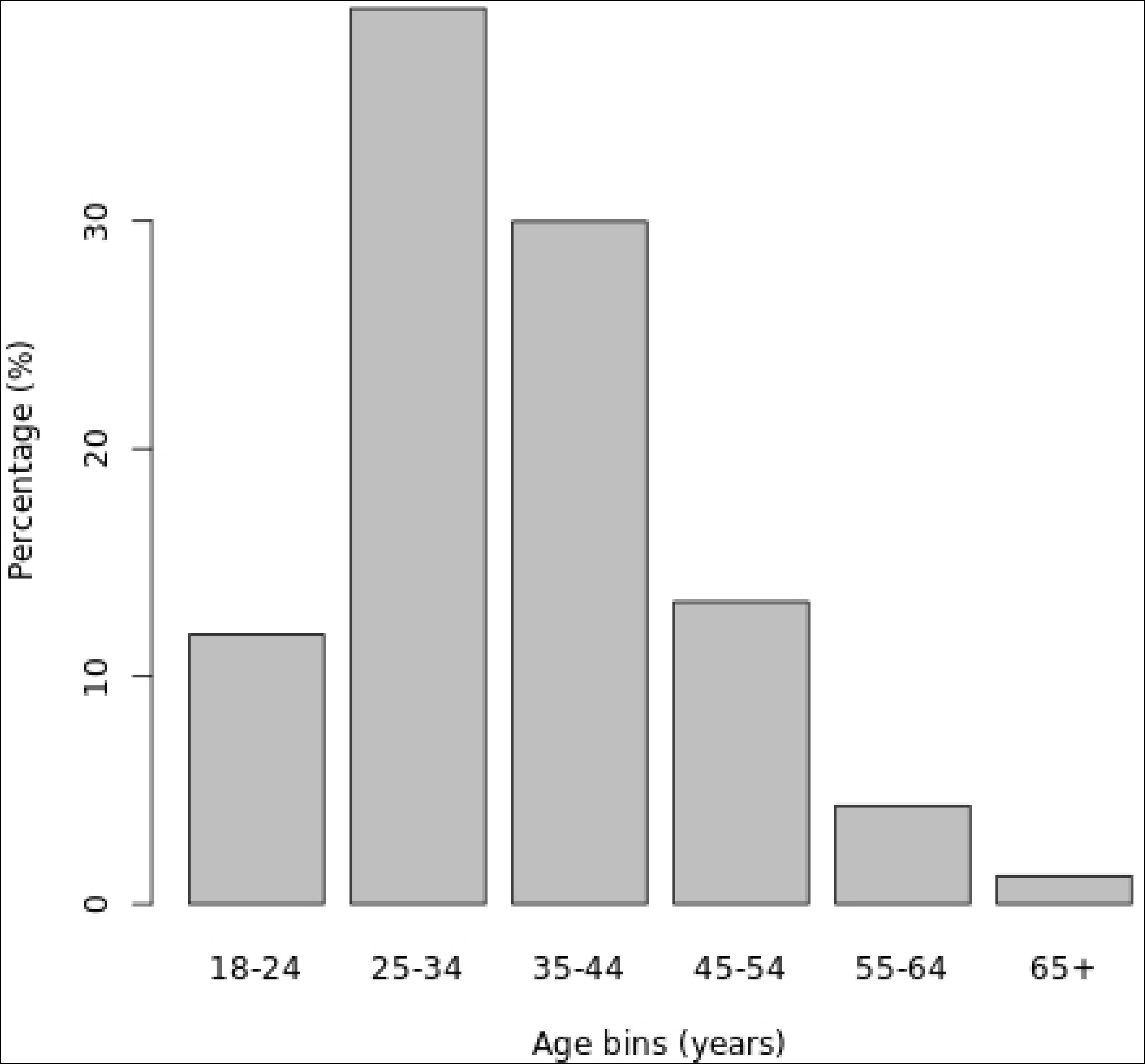
Distribution of age categories.

During the study period, 500,729 (54.10%) patients experienced an IIT. Within the first month of care, 0.1% experienced their first interruption; within three months, this increased to 29.28%, and this further increased to 46.49% in the first six months and 50.26% in the first year. The distribution of participant characteristics between censored and interrupted observations is presented in Table 1. A higher proportion of women experienced interruptions, with 345,115 (56.93%) having an interruption compared to 155,614 (48.84%) males. Age was also associated with interruptions (p < 0.001). The proportion of interruptions increased progressively with increasing age categories 55,454 (50.16%) in the 18-24 years group and was highest in the 55-64 years group with 23,830 (50.58%) interruptions. The number of people aged 65 years and older who experienced IIT was 6,752 (59.11%).

**Table 1:** Variable characteristics in people who presented for a clinic visit.

|  | No Interruption in Treatment<br>(N/%) | Interruption in Treatment<br>(N/%) | P-value |
| --- | --- | --- | --- |
| **Sex** |  |  | <0.001 |
| **Women** | 261,107 (43.07%) | 345,115 (56.93%) |  |
| **Men** | 163,011 (38.4%) | 155,614 (48.84%) |  |
| **Age Categories** |  |  | <0.001 |
| **18-24years** | 54,112 (49.39%) | 55,454 (50.61%) |  |
| **25-34years** | 172,416 (47.39%) | 191,383 (52.61%) |  |
| **35-44years** | 124,174 (44.78%) | 153,107 (55.22%) |  |
| **45-54years** | 52,580 (42.82%) | 70,203 (57.18%) |  |
| **55-64years** | 16,165 (40.42%) | 23,830 (59.58%) |  |
| **65+ years** | 4,671 (40.89%) | 6,752 (59.11%) |  |
| **Anti retroviral therapy** |  |  | <0.001 |
| **INSTI** | 284,802(65.58%) | 149,487 (29.90%) |  |
| **NNRTI** | 70,468 (19.05%) | 299,423 (80.95%) |  |
| **PI** | 1,013 (18.01%) | 4,611 (81.99%) |  |
| **Undocumented** | 67,835 (58.96%) | 47,208 (41.04%) |  |
| **CD4 recorded** |  |  | <0.001 |
| **No** | 378,398 (53.34%) | 331,060 (46.66%) |  |
| **Yes** | 45,720 (21.23%) | 169,669 (78.77%) |  |
| **HIV Viral Load recorded** |  |  | <0.001 |
| **No** | 347,586 (57.20%) | 260,066 (42.80%) |  |
| **Yes** | 76,532 (24.13%) | 240,663 (75.87%) |  |
| **WHO Clinical Stage** |  |  | <0.001 |
| **Unknown** | 19,368 (41.23%) | 27,610 (58.77%) |  |
| **Stage 1** | 369,282 (49:17%) | 381,787 (50.83%) |  |
| **Stage 2** | 20,651 (26.03%) | 58,678 (73.97%) |  |
| **Stage 3** | 13,560 (30.81%) | 30,448 (69.19%) |  |
| **Stage 4** | 1,257 (36.30%) | 2,206 (63.70%) |  |
| **Functional Status** |  |  | <0.001 |
| **Unknown** | 14,409 (31.99%) | 30,632 (68.01%) |  |
| **Bedridden** | 1,082 (53.62%) | 936 (46.38%) |  |
| **Ambulatory** | 2,278 (33.09%) | 4,607 (66.91%) |  |
| **Working** | 406,349 (46.66%) | 464,554 (53.34%) |  |
| **Last measured weight\*** | 58 (18) | 57 (19) | <0.001 |
| **Highest education attained** |  |  | <0.001 |
| **Unknown** | 198,716 (41.40%) | 281,226 (58.60%) |  |
| **None** | 29,329 (47.49%) | 32,429 (52.51%) |  |
| **Q'ranic** | 6,692 (33.79%) | 13,115 (66.21%) |  |
| **Primary** | 41,198 (51.06%) | 39,492 (48.94%) |  |
| **Secondary** | 120,869 (54.43%) | 101,210 (45.57%) |  |
| **Tertiary** | 27,314 (45.09%) | 33,257 (54.91%) |  |
| **Occupation** |  |  | <0.001 |
| **Unknown** | 197,033 (42.52%) | 266,355 (57.48%) |  |
| **Unemployed** | 127,083 (50.02%) | 126,991 (49.98%) |  |
| **Student** | 14,035 (52.15%) | 12,878 (47.85%) |  |
| **Employed** | 85,243 (8.37%) | 93,403 (91.63%) |  |
| **Retired** | 724 (39.65%) | 1,102 (60.35%) |  |
| **Marital Status** |  |  | <0.001 |
| **Unknown** | 200,166 (41.71%) | 279,713 (58.29%) |  |
| **Single** | 109,423 (58.63%) | 77,221 (41.37%) |  |
| **Married** | 114,529 (44.34%) | 143,794 (55.66%) |  |
| **Other** | 0 (0.00%) | 1 (100.00%) |  |
| **Year enrolled in care** |  |  | <0.001 |
| **2015** | 15,399 (17.20%) | 74,136 (82.80%) |  |
| **2016** | 17,693 (16.58%) | 89,044 (83.42%) |  |
| **2017** | 19,866 (17.60%) | 93,006 (82.40%) |  |
| **2018** | 25,291 (23.95%) | 80,295 (76.05%) |  |
| **2019** | 55,781 (37.59%) | 92,601 (62.41%) |  |
| **2020** | 199,897 (74.10%) | 69,880 (25.90%) |  |
| **2021** | 90,191 (98.08%) | 1,767 (1.92%) |  |
| **Enrolled after Surge commencement** |  |  | <0.001 |
| **No** | 78,249 (18.87%) | 336,481 (81.13%) |  |
| **Yes** | 345,869 (67.80%) | 164,248 (32.20%) |  |
| **Enrolled after COVID** |  |  | <0.001 |
| **No** | 75,351 (16.63%) | 377,656 (83.37%) |  |
| **Yes** | 348,767 (73.92%) | 123,073 (26.08%) |  |
| **Urban vs Rural** |  |  | <0.001 |
| **Urban dweller** | 340,328 (80.2%) | 415,230 (82.9%) |  |
| **Rural dweller** | 83,790 (19.8%) | 85,499 (17.1%) |  |
| **Hospital Encounters\*** | 3 (2,5) | 2 (2,4) | <0.001 |
| **Time to interruption event**<br>**(months)\*** | 7 (3,12) | 9 (5,16) | <0.001 |

\*Median(Interquartile range) reported
*NNRTI: Non nucleoside reverse transcriptase inhibitor*
*PI: Protease inhibitor*
*INST: Integrase strand transfer inhibitors*

Non-NRTI drugs in the antiretroviral therapy (ART) regimen were associated with interruptions (p-value < 0.001). Among the different regimens, individuals on the INSTI had the highest proportion of retention in care (Fig 2), with 284,803 (65.58%). The NNRTIs had the highest interruption rate of 299,423(80.95%). PIs had 4,611(81.99%) interruptions, while undocumented regimens had 67,835(58.96%) interruptions. Those with recorded CD4 levels had a higher proportion of interruptions (169,669; 78.77%) compared to 46.66% for those with no records of CD4 count, similarly, those with recorded viral load had higher interruptions (240,663; 75.87%) compared to 42.80% for those with no recorded HIV viral load. The proportion with ambulatory functional status was most likely to experience interruption with 6.885(66.91%) experiencing interruption when compared to those who were bedridden (936,(46.38%)) or working (464554,(53.34%)).

**Fig 2.**
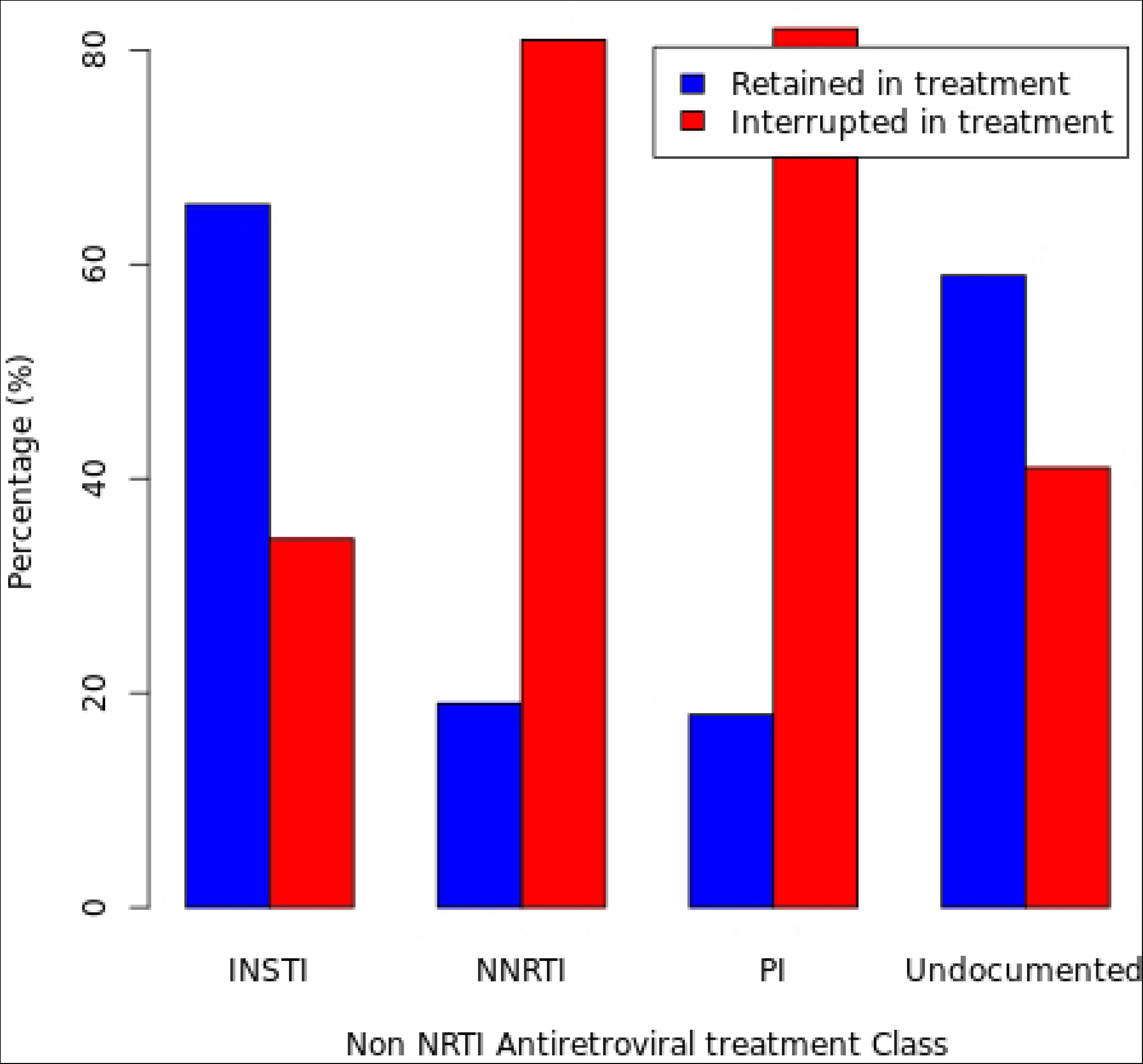
Distribution of Time to first IIT event by Antiretroviral medication.

PLHIV with primary and secondary education had lower proportions of interruption: 39,492(48.94%) and 101,210(45.57%), respectively while no education, qur’anic, and tertiary education had higher proportions of interruption: 32,429(52.51), 13,115(66.21%), and 33,257(54.91%), respectively. Employed people had the highest proportion of interruptions compared with other occupation groups, with 933,403(91.63%) experiencing interruption. This was followed by 1,102(60.35%) retirees. Students and unemployed had lower proportions of interruptions with 12,878(47.85%) and 126,991(49.98%), respectively.

Those enrolled in 2015, 2016, and 2017 had a higher proportion of interruptions, with 74,136 (82.80%), 89,044 (83.42%), and 93,006 (82.40%) interruptions, respectively. A higher proportion of interruptions occurred in patients who enrolled pre-surge (336,481, 81.13%) and pre-COVID (75,351; 83.87%).

The median number of hospital encounters differed between those who continued in care (3, IQR = (2,5)) and those interrupted (2, IQR = (2,4)) observations (p < 0.001). On average patients with interrupted observations had fewer hospital visits. Patients with interrupted observations experienced a longer time to interruption (median: 7 months; IQR: (3,9)) event than those who continued receiving ART uninterrupted (median: 9 months, IQR:(5,16)) without interruption.

### Logistic regression of interruption to treatment

The results of the statistical analysis are presented in Table 2; sex was associated with the outcome, with men having 0.90 (0.89, 0.91) times lower odds of IIT compared to women. Compared to people aged 25-34 years, there were no differences in the odds of experiencing IIT for those aged 18-24 years (aOR = 0.98, 95% CI: 0.96-1.00, p = 0.074), 35-44 years (aOR = 1.03, 95% CI: 1.01-1.04, p = 0.002), 45-54 years (aOR = 1.02, 95% CI: 0.99-1.04, p = 0.136), and 55-64 years (aOR = 1.02, 95% CI: 0.98-1.04, p = 0.321) and in the 65 years and above group (aOR = 0.96, 95% CI: 0.90-1.02, p = 0.147).

**Table 2:** Factors associated with interruption in HIV treatment among people living with HIV in Nigeria, 2015-2021.

|  | Adjusted Odds Ratios (aOR) | 95% Confidence Interval | p-value |
| --- | --- | --- | --- |
| **Male sex (Ref. Women)** | 0.90 | 0.89 – 0.91 | <0.001 |
| **Age Category (Ref. 25-34years)** |  |  |  |
| 18-24 years | 0.98 | 0.96 – 1.00 | 0.074 |
| 35-44 years | 1.03 | 1.01 – 1.04 | 0.002 |
| 45-54 years | 1.02 | 0.99 – 1.04 | 0.136 |
| 55-64 years | 1.02 | 0.98 – 1.05 | 0.321 |
| 65+ years | 0.96 | 0.90 – 1.02 | 0.147 |
| **ART Regimen (Ref. INSTI)** |  |  |  |
| **NNRTI** | 1.33 | 1.30 – 1.36 | <0.001 |
| **PI** | 1.09 | 0.97 – 1.23 | 0.161 |
| **Undocumented** | 1.02 | 0.99 – 1.06 | <0.001 |
| **Viral Load Recorded (Ref: Not recorded)** | 3.12 | 3.07 – 3.16 | <0.001 |
| **CD4 Recorded (Ref. Not recorded)** | 1.03 | 1.01 – 1.05 | <0.001 |
| **WHO clinical staging (Ref. Stage 1)** |  |  |  |
| **Stage 2** | 0.99 | 0.97 – 1.02 | <0.001 |
| **Stage 3** | 0.55 | 0.54 – 0.57 | <0.001 |
| **Stage 4** | 0.39 | 0.35 – 0.43 | <0.001 |
| **Undocumented** | -- | -- | -- |
| **Functional Status (Ref. Working)** |  |  |  |
| **Ambulatory** | 0.36 | 0.35 – 0.38 | <0.001 |
| **Bedridden** | 0.49 | 0.42 – 0.57 | <0.001 |
| **Unknown functional status** | 0.59 | 0.55 – 0.63 | <0.001 |
| **Last measured Weight** | 3.12 | 3.07 – 3.16 | <0.001 |
| **Highest Education (Ref. Secondary)** |  |  |  |
| **No education** | 1.01 | 0.97 – 1.04 | 0.724 |
| **Qur'anic** | 0.90 | 0.88 – 0.93 | <0.001 |
| **Primary** | 0.97 | 0.92 – 1.02 | 0.242 |
| **Tertiary** | 0.82 | 0.79 – 0.84 | <0.001 |
| **Unknown** | 1.04 | 1.01 – 1.07 | 0.017 |
| **Occupation (Ref. Employed)** |  |  |  |
| **Retired** | 0.93 | 0.90 – 0.96 | <0.001 |
| **Student** | 1.12 | 1.07 – 1.17 | <0.001 |
| **Unemployed** | 0.94 | 0.91 – 0.97 | <0.001 |
| **Unknown** | 0.91 | 0.79 – 1.05 | 0.206 |
| **Marital Status (Ref. Married)** |  |  |  |
| **Single** | 0.96 | 0.93 – 0.98 | 0.001 |
| **Unknown** | -- | -- | -- |
| **Year Enrolled** | 0.70 | 0.69 – 0.70 | <0.001 |
| **Surge Enrollee(Ref. Pre Surge enrollee)** | 1.48 | 1.43 – 1.52 | <0.001 |
| **COVID Enrollee (Ref. Pre COVID enrollee)** | 0.19 | 0.19 – 0.19 | <0.001 |
| **Urban dweller (Ref. Rural dweller)** | 0.83 | 0.80 – 0.85 | <0.001 |

*NNRTI: Non nucleoside reverse transcriptase inhibitor*
*PI: Protease inhibitor*
*INST: Integrase strand transfer inhibitors*

The type of non-NRTI drug used in the antiretroviral (ART) regimen was associated with IIT. Compared to individuals on the INSTI regimen, those on NNRTI regimen had higher odds of the outcome (aOR = 1.33, 95% CI: 1.30-1.36, p = <0.001. Individuals on PI and undocumented regimens showed no significant difference in adjusted odds ratios of 1.09 (0.97, 1.23) and 1.02 (0.99, 1.06) respectively. People with a recorded viral load had higher odds of the outcome compared to those without a recorded viral load (aOR = 3.12, 95% CI: 3.07-3.16, p < 0.001) as with, to a lesser degree, people with recorded CD4 levels (aOR = 1.03, 95% CI: 1.01-1.05, p < 0.001).

People with a WHO Stage of II(aOR = 0.99, 95% CI: 0.97-1.02, p < 0.001), Stage III (aOR = 0.55, 95% CI: 0.54-0.57, p < 0.001), and Stage IV (aOR = 0.39, 95% CI: 0.35-0.43, p < 0.001) had lower odds of IIT compared to those in Stage I. Being ambulatory (aOR = 0.36, 95% CI: 0.35-0.38, p < 0.001) or bedridden (aOR = 0.49, 95% CI: 0.42-0.57, p < 0.001) had lower odds of the outcome compared to those classified as working.

Using secondary school education as a reference, no education (aOR = 1.01, 95% CI: 0.97-1.04, p = 0.724), primary education (aOR = 0.97, 95% CI: 0.92-1.02, p = 0.242) or undocumented education (aOR = 1.04, 95% CI: 1.01-1.07, p = 0.017) were not associated with IIT. Qur’anic education (aOR = 0.90, 95% CI: 0.88-0.93, p < 0.001), and tertiary education (aOR = 0.82, 95% CI: 0.79-0.84, p < 0.001) were associated with lower odds of the outcome. Retired (aOR = 0.93, 95% CI: 0.90-0.96, p < 0.001), and unemployed (aOR = 0.94, 95% CI: 0.91-0.97, p < 0.001) individuals had lower odds of the outcome compared to employed individuals. Students (aOR = 1.12, 95% CI: 1.07-1.17, p < 0.001) had higher odds of IIT compared to employed individuals. Having undocumented education was not associated with IIT (aOR=0.91, 95%CI:0.79-1.05). Being single was protective against IIT (aOR = 0.96, 95% CI: 0.93-0.98, p < 0.001) compared to those who were married.

The year PLHIV were enrolled in treatment was associated with the outcome, with lower odds of the outcome for more recent enrollments (aOR = 0.70, 95% CI: 0.69-0.70, p < 0.001). Starting ART during the national surge efforts was associated with higher odds of IIT compared to the pre-surge period (aOR = 1.48, 95% CI: 1.43-1.52, p < 0.001), and enrollment during the early phase of the COVID-19 pandemic was associated with lower odds of IIT compared to the pre-COVID-19 period (aOR = 0.19, 95% CI: 0.19-0.19, p < 0.001). Residing in an urban area was associated with lower odds of the outcome than residing in a rural area (aOR = 0.83, 95% CI: 0.80-0.85, p < 0.001).

### Survival analysis of the first interruption in care of PLHIV

The initial exploration of the categorical variables using a long rank test and Kaplan-Meier plots (Fig 3). All categorical variables were statistically different in the life test with p-values <0.001. The results from the multivariable analysis using Cox proportional hazards regression are presented in Table 3, showing the adjusted hazard ratios (aHR) along with their associated 95% confidence intervals (CI) and p-values for each variable. Sex was not associated with the outcome, with men having an adjusted hazard ratio of 1.00 (95% CI: 0.99-1.01) compared to women (p = 0.938). PLHIV aged 18-24 years had a 1.16 (1.14, 1.17) times higher hazard of the outcome compared to those aged 25-34 years. Conversely, those aged 35-44 years (aHR = 0.92, 95% CI: 0.91-0.93, p < 0.001), 45-54 years (aHR = 0.91, 95% CI: 0.90-0.92, p < 0.001), 55-64 years (aHR = 0.89, 95% CI: 0.88-0.90, p < 0.001), and 65 years and above (aHR = 0.89, 95% CI: 0.86-0.92, p < 0.001) had lower hazards of the outcome.

**Fig 3.**
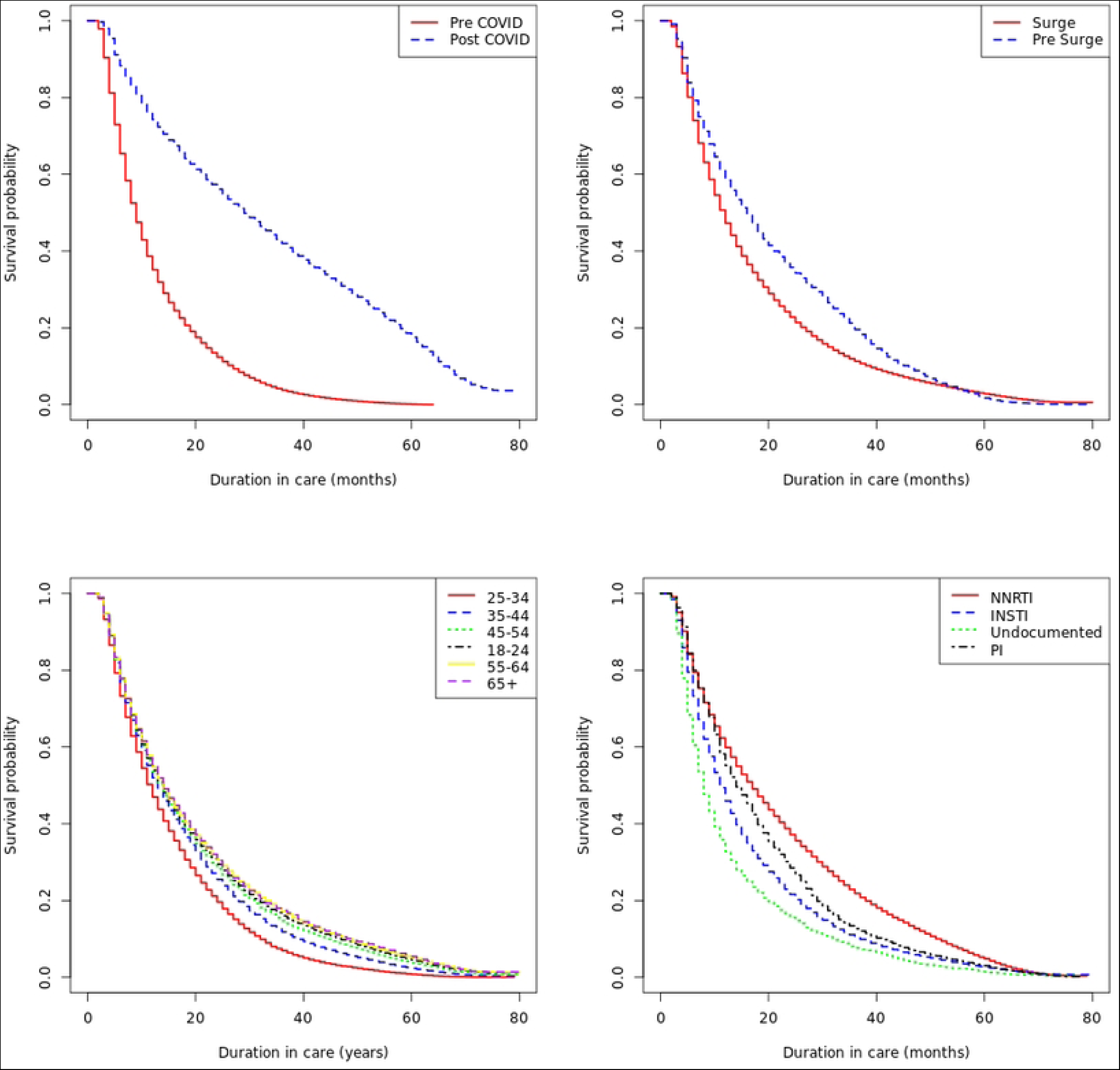
Kaplan Meier plots showing time to first interruption in ART versus enrollment pre/post surge commencement; pre/post-COVID pandemic; Antiretroviral therapy; and Age categories.

**Table 3:** Factors associated with time to first interruption in HIV treatment among people living with HIV in Nigeria, 2015-2021.

|  | Adjusted<br>Hazard Ratios<br>(aHR) | 95 % Confidence<br>Interval | P-value |
| --- | --- | --- | --- |
| **Male sex (Ref: Women)** | 1.00 | 0.99 – 1.01 | 0.938 |
| **Age Category (Ref. 25-34years)** |  |  |  |
| **18-24 years** | 1.16 | 1.14 – 1.17 | <0.001 |
| **35-44 years** | 0.92 | 0.91 – 0.93 | <0.001 |
| **45-54 years** | 0.91 | 0.90 – 0.92 | <0.001 |
| **55-64 years** | 0.89 | 0.88 – 0.90 | <0.001 |
| **65+ years** | 0.89 | 0.86 – 0.92 | <0.001 |
| **ART Regimen (Ref. INSTI)** |  |  |  |
| **NNRTI** | 0.80 | 0.80 – 0.81 | <0.001 |
| **PI** | 1.07 | 1.03 – 1.11 | <0.001 |
| **Undocumented** | 0.90 | 0.88 – 0.92 | <0.001 |
| **Viral Load Recorded (Ref: Not recorded)** | 1.05 | 1.04 – 1.05 | <0.001 |
| **CD4 Recorded (Ref. Not recorded)** | 0.89 | 0.88 – 0.89 | <0.001 |
| **WHO clinical staging (Ref. Stage 1)** |  |  |  |
| **Stage 2** | 1.19 | 1.18 – 1.20 | <0.001 |
| **Stage 3** | 1.27 | 1.26 – 1.29 | <0.001 |
| **Stage 4** | 1.26 | 1.20 – 1.33 | <0.001 |
| **Missing** | 1.31 | 1.29 – 1.34 | <0.001 |
| **Functional Status (Ref. Working)** |  |  |  |
| **Ambulatory** | 0.90 | 0.88 – 0.92 | <0.001 |
| **Bedridden** | 1.09 | 1.00 – 1.19 | 0.040 |
| **Unknown functional status** | 1.09 | 1.05 – 1.13 | <0.001 |
| **Last measured Weight** | 1.00 | 1.00 – 1.00 | <0.001 |
| **Highest Education (Ref. Secondary)** |  |  |  |
| **No education** | 1.00 | 0.99 – 1.02 | 0.963 |
| **Q'ranic** | 0.98 | 0.97 – 0.99 | 0.001 |
| **Primary** | 1.01 | 0.98 – 1.03 | 0.666 |
| **Tertiary** | 0.92 | 0.91 – 0.94 | <0.001 |
| **Unknown** | 0.99 | 0.98 – 1.01 | 0.311 |
| **Occupation (Ref. Employed)** |  |  |  |
| **Retired** | 1.01 | 0.99 – 1.02 | 0.351 |
| **Student** | 1.01 | 0.98 – 1.03 | 0.625 |
| **Unemployed** | 1.04 | 1.03 – 1.06 | 0.005 |
| **Unknown** | 1.10 | 1.03 – 1.17 | 0.012 |
| **Marital Status (Ref. Married)** |  |  |  |
| **Single** | 0.98 | 0.97 – 1.00 | 0.007 |
| **Unknown** | -- | -- | -- |
| **Year Enrolled** | 1.21 | 1.20 – 1.21 | <0.001 |
| **Surge Enrollee(Ref. Pre Surge enrollee)** | 1.14 | 1.12 – 1.15 | <0.001 |
| **COVID Enrollee (Ref. Pre COVID enrollee)** | 0.17 | 0.17 – 0.17 | <0.001 |
| **Urban dweller (Ref. Rural dweller)** | 1.06 | 1.05 – 1.07 | <0.001 |

*NNRTI: Non nucleoside reverse transcriptase inhibitor*
*PI: Protease inhibitor*
*INST: Integrase strand transfer inhibitors*

Compared to individuals on the INSTI regimen, those on NNRTI (aHR = 0.80, 95% CI: 0.80-0.81, p < 0.001), and undocumented (aHR = 0.90, 95% CI: 0.88-0.92, p < 0.001) had a lower risk of interruptions in treatment within the same time period. Those on PI (aHR = 1.07, 95% CI: 1.03-1.11, p < 0.001) had a higher risk for interruption within the same time period when compared to those on INSTI. Patients with a recorded viral load had a 1.05 (1.04, 1.05) times higher instantaneous risk of the outcome compared to those without a recorded viral load. Those with recorded CD4 levels had a lower risk 0.89 (0.88, 0.89) than those without recorded CD4 levels within the same time period.

Individuals with Stage II (aHR = 1.19, 95% CI: 1.18-1.20, p < 0.001), Stage III (aHR = 1.27, 95% CI: 1.26-1.29, p < 0.001), Stage IV (aHR = 1.26, 95% CI: 1.20-1.33, p < 0.001), and those with missing staging information (aHR = 1.31, 95% CI: 1.29-1.34, p < 0.001) had a higher risk of first interruption within the same time period compared to those in Stage I. Ambulatory patients (aHR = 0.90, 95% CI: 0.88-0.92, p < 0.001) had a lower instantaneous risk of the outcome, while being bedridden (aHR = 1.09, 95% CI: 1.00-1.19, p = 0.040) was not associated with the outcome. Those with unknown functional status (aHR = 1.00, 95% CI: 1.00-1.00, p < 0.001) were not associated with the outcome.

Being uneducated (aHR = 0.1.00, 95% CI: 0.99-1.02, p < 0.001), having primary education (aHR = 1.01, 95%CI: 0.98-1.03), and undocumented education (aHR = 0.99, 95%CI: 0.98-1.01) were not associated with instantaneous risk of first interruption in treatment. Qur’anic education (aHR = 0.98, 95% CI: 0.97-0.99, p = 0.001), and tertiary education (aHR = 0.92, 95%CI: 0.91-0.94) were associated with a lower instantaneous risk of being interrupted compared to secondary education. When comparing being employed with retired (aHR = 1.01, 95% CI: 0.99-1.02), a student (aHR = 1.01, 95% CI: 0.98-1.03), unemployed (aHR = 1.04, 95%CI: 1.03-1.06), and undocumented occupation (aHR = 1.10, 95%CI: 1.03-1.17) did not show any association. Being single was not associated with an instantaneous risk of treatment interruption when compared to being married (aHR = 0.98, 95% CI: 0.97-1.00, p = 0.007).

The year of enrollment in the study was associated with the outcome, with higher hazards for more recent enrollments (aHR = 1.21, 95% CI: 1.20-1.21, p < 0.001). Being a Surge Enrollee (aHR = 1.14, 95% CI: 1.12-1.15, p < 0.001) and being a COVID Enrollee (aHR = 0.17, 95% CI: 0.17-0.17, p < 0.001) remained significantly associated with different hazards of the outcome compared with being a pre Surge or Pre COVID enrollee, respectively. Residing in an urban area showed an association with the outcome, with individuals from urban areas having a 1.06 (95% CI: 1.05-1.07) times higher risk of being interrupted in treatment within the same time period than those from rural areas (p < 0.001).

## Discussion

In this study, we investigated the predictors of treatment interruption using data from the national HIV treatment program. We identified sets of predictors of first interruption in treatment using the logistic regression and these to be consistent in predicting time to first interruption including sex, non NRTI drug in ART regimen, recorded HIV viral load, recorded CD4 cell count, WHO clinical staging, functional status, last measured weight, highest education attained, occupation, marital status, year enrolled in care, pre and post surge, pre and post-COVID and residing in a state capital, Lagos, FCT (urban) versus other locations (rural). Age grouping was the only variable that was predictive only for time to first interruption but not for having a first interruption.

Logistic regression captured some predictors of IIT, including female sex. In the 2019 study by Aliyu et al[17] 2021 study by Tomescu et al[16], the 2022 study by Hakizayezu et al[13], and a 2020 study by Abebe et al[20] sex was found to be predictive of interruptions in care; however, in all of these studies, the men were at a higher risk of interruption or its proxy such as loss to follow up. Our findings may differ from those of prior studies because we were restricted to those who interrupted but re-engaged in care. It may be that men are more likely to discontinue care completely and that women interrupt and return. This then requires more intense follow-up or development of new strategies to keep men in care. PLHIV at WHO clinical stage I for AIDS had the highest odds of experiencing IIT (this is also at variance with the Abebe et al study where Stages I and II combined were demonstrated to have a lower risk of IIT[20]). PLHIV with the functional status of working were most likely to experience IIT (the reverse was seen in the Abebe et al study with ambulatory or bedridden at greater risk[20]). This pattern may suggest that people who feel they are healthy are more likely to interrupt and consequently drop out of care hence the need for educating PLHIV that as their status improves they need to remain in care as well as encouraging those without symptoms to remain in care and educating them on the risks of interruption. Students when compared with the employed were at higher risk of interruption. We found that being retired or unemployed was more protective than employment. Being uneducated was identified as a risk factor, consistent with the Abebe et al study that compared unemployment to employment. The enrollment year variable was suggestive of the longer people were in care the more likely they were to experience an interruption event. This is consistent with the finding in the Aliyu et al study where duration on ART was found to be related to the odds of loss to follow up with the odds of remaining in care improving with the longer duration of treatment[17]. PLHIV enrolled after the commencement of the surge effort were at a higher risk of IIT whereas PLHIV enrolled before COVID were at a higher risk of experiencing IIT. People living outside of state capitals, the federal capital territory, and Lagos State were more likely to experience at least one interruption event. The Hakizayezu et al study identified other predictors, including a prior history of treatment interruption (which we could not explore as were looking at first interruption) and good perception toward the whole-of-life treatment[13]. Aliyu et al identified additional predictors including HIV viral load indication and result as well as age[17] (which was not predictive in this study or in the Tomescu et al study[16]), and regimen at the start of ART. Other identified predictors in the Tomescu et al study included geo-political zone, regimen line, and viral load suppression[16].

The findings with the proportional hazards regression were in some cases as expected as opposed to those of the logistic regression. Sex was not predictive of time to treatment interruption unlike in predicting being interrupted. The instantaneous risk worsened as the WHO clinical stage worsened in our study when compared to stage I. Age groups, which was not predictive of being interrupted in treatment, were predictive of time to interruption with a trend towards a lower risk with increasing age. This was consistent with the APIN study where younger age was found to be predictive of people under 30 years of age at the greatest risk of LTFU[14]. We found that the ART class of medication was predictive with NNRTI, and PI having greater instantaneous risk than INSTI. This is contrary to the 2014 Meloni et al study that did not find starting an ART regimen to be predictive[14]. Being married had a higher instantaneous risk of IIT than being single contrary to the reverse that was found in the Meloni et al study in Nigeira[14]. Having HIV viral load measured posed greater instantaneous risk in the case of logistic regression however the reverse was found for having measured and recorded CD4 cell count. In most of the time periods covered in the dataset, CD4 was not required for ART initiation and was run for cause (regimen choice, etc.). In addition, HIV viral load was not run at baseline and, as such, likely is a pointer to engagement in care. Consistent with logistic regression a working functional status had a higher instantaneous risk than ambulatory status. Having Q’ranic or tertiary education were both demonstrated to be protective of IIT when compared to secondary education, and this is consistent with the Meloni et al study where tertiary education was found to be protective compared to secondary education[14]. For the year of enrolment, we found the instantaneous risk increases as the years become more recent (moving from 2015 to 2021), again in contrast to the findings with the logistic model. The Meloni et al study did not find a significant difference in the year of ART initiation[14]. However consistent with logistic regression we find that people who enrolled after the commencement of the surge effort are more likely to experience interruption while those enrolled after COVID are protected. Urban dwellers (in state capitals, FCT, or Lagos) have a higher instantaneous risk of interruption. In the Meloni et al study, additional predictors identified included CD4 cell count[11], [14] while in the NCH study baseline CD4 count was found to be a predictor[11]. Across the two approaches used, we found some predictors to have intersected including measured and recorded HIV viral load, surge enrollment, and COVID enrollment. Others that were predictive but in different directions included sex, measured and recorded CD4 count, marital status, and WHO clinical stage.

The rate of missed appointments increased as the duration of care progressed, emphasizing the need for interventions to improve appointment adherence, not only in the early stages of care but also as treatment fatigue potentially sets in[21]. The 25-34 age group constituted the largest proportion of the population, indicating the importance of targeted interventions and support for this age cohort who are also at higher risk of interruption in treatment. We also found weight change to be influential, with weight gain associated with an increased risk of interruption. Importantly, the COVID-19 pandemic had a significant impact on interruptions in care. PLHIV enrolled after the outbreak demonstrated a lower risk of interruption, potentially due to increased emphasis on healthcare access and continuity during the pandemic. However, individuals enrolled before the pandemic experienced a higher risk of interruption, indicating the need for targeted interventions to mitigate the effects of external factors on HIV care. Enrollment after the initiation of the surge initiative was linked to a higher risk of interruption which begs the question of the quality of follow-up after aggressive identification of cases and enrollment into care. Overall, the findings of this study contribute to our understanding of care interruptions. A huge takeaway from this study is the potential for differentiated service delivery, a person-centric approach to care and treatment in HIV in Nigeria and sub-Saharan Africa as we see multiple factors interacting to determine the outcomes of interruption in treatment and progression to loss to follow-up.

### Limitations of the Study

Our study design was retrospective, limiting our control over data collection and the availability of variables of interest including access to care and type of healthcare facility. In addition, we did not include certain variables because of the degree of missingness such as CD4 cell count, HIV viral load (both of which were dichotomized to included or missing), pregnancy status, comorbidities including tuberculosis, and other potential confounders. The findings of this study are dependent on the accuracy and completeness of the recorded data. Potential limitations such as missing or incomplete data, coding errors, or inaccuracies in medical records could affect the validity of the results.

## Conclusion

To reduce the risk of IIT it is important to target interventions preemptively. We have highlighted the need for tailored interventions that address the unique needs of PLHIV in Nigeria. Targeted interventions focusing on those with a combination of risk factors could include education, counseling, supportive services, and monitoring and outreach. Education and counseling are critical in addressing the challenges of treatment adherence. There should be continued education, and improvement on education strategies for PLHIV on the importance of adhering to treatment and the risks of IIT augmented with appointment reminders and rescheduling[22]. Supportive services such as case management and peer support groups can help PLHIV to stay on track with their treatment and to manage any challenges they may face[22]. Monitoring and outreach are essential in identifying PLHIV who are at high risk for IIT[22]. By closely monitoring PLHIV closely, healthcare providers can identify signs of IIT and provide support promptly. Outreach to PLHIV who are at risk of missing appointments can be helpful as a part of providing support to stay in care[22].

## Data Availability

Data cannot be shared publicly because it is the property of the Nigerian Federal MInistry of Health. Data are available from the Nigerian Federal Ministry of Health (contact via email) for researchers who meet the criteria for access to confidential data.

https://ndr.phis3project.org.ng/

## Acknowledgments

We would like to acknowledge the Nigerian Federal Ministry of Health for providing the data from the National Data Repository for this study. The data from the National Data Repository were essential for conducting this study and we are grateful for the opportunity to have access to this valuable resource.

## Supporting Information

**Table 4:** Characteristics of Month of interruption and state (location) by interruption status.

|  | No interruption in treatment | Interruption in treatment | p-value |
| --- | --- | --- | --- |
| Month of interruption |  |  |  |
| January | 10865 (2.6%) | 50287 (10.0%) | <0.001 |
| February | 29541 (7.0%) | 45649 (9.1%) |  |
| March | 42279 (10.0%) | 50070 (10.0%) |  |
| April | 52780 (12.4%) | 34052 (6.8%) |  |
| May | 90575 (21.4%) | 35381 (7.1%) |  |
| June | 88122 (20.8%) | 39169 (7.8%) |  |
| July | 56814 (13.4%) | 35408 (7.1%) |  |
| **August** | 15981 (3.8%) | 39425 (7.9%) |  |
| **September** | 9949 (2.3%) | 41106 (8.2%) |  |
| **October** | 9766 (2.3%) | 40996 (8.2%) |  |
| **November** | 8951 (2.1%) | 45980 (9.2%) |  |
| **December** | 8495 (2.0%) | 43206 (8.6%) |  |
| **State** |  |  |  |
| **Abia** | 1758 (0.4%) | 1678 (0.3%) | <0.001 |
| **Adamawa** | 7243 (1.7%) | 12490 (2.5%) |  |
| **Akwa Ibom** | 82318 (19.4%) | 54054 (10.8%) |  |
| **Anambra** | 4692 (1.1%) | 13244 (2.6%) |  |
| **Bauchi** | 6172 (1.5%) | 9539 (1.9%) |  |
| **Bayelsa** | 4271 (1.0%) | 4026 (0.8%) |  |
| **Benue** | 32416 (7.6%) | 31135 (6.2%) |  |
| **Borno** | 3028 (0.7%) | 5533 (1.1%) |  |
| **Cross River** | 16588 (3.9%) | 24123 (4.8%) |  |
| **Delta** | 21473 (5.1%) | 9217 (1.8%) |  |
| **Ebonyi** | 960 (0.2%) | 898 (0.2%) |  |
| **Edo** | 3224 (0.8%) | 8567 (1.7%) |  |
| **Ekiti** | 563 (0.1%) | 356 (0.1%) |  |
| **Enugu** | 7478 (1.8%) | 3744 (0.7%) |  |
| **FCT** | 22395 (5.3%) | 14832 (3.0%) |  |
| **Gombe** | 3605 (0.9%) | 4104 (0.8%) |  |
| **Imo** | 4878 (1.2%) | 6224 (1.2%) |  |
| **Jigawa** | 1457 (0.3%) | 3453 (0.7%) |  |
| **Kaduna** | 4393 (1.0%) | 4614 (0.9%) |  |
| **Kano** | 4217 (1.0%) | 12724 (2.5%) |  |
| **Katsina** | 1918 (0.5%) | 2465 (0.5%) |  |
| **Kebbi** | 2940 (0.7%) | 4882 (1.0%) |  |
| **Kogi** | 3655 (0.9%) | 4760 (1.0%) |  |
| **Kwara** | 1589 (0.4%) | 3008 (0.6%) |  |
| **Lagos** | 34119 (8.0%) | 34517 (6.9%) |  |
| **Nasarawa** | 12254 (2.9%) | 11043 (2.2%) |  |
| **Niger** | 6481 (1.5%) | 10698 (2.1%) |  |
| **Not recorded** | 47950 (11.3%) | 123001 (24.6%) |  |
| **Ogun** | 3702 (0.9%) | 7145 (1.4%) |  |
| **Ondo** | 1856 (0.4%) | 1443 (0.3%) |  |
| **Osun** | 1068 (0.3%) | 1187 (0.2%) |  |
| **Oyo** | 2750 (0.6%) | 7664 (1.5%) |  |
| **Plateau** | 3222 (0.8%) | 7092 (1.4%) |  |
| **Rivers** | 58358 (13.8%) | 38431 (7.7%) |  |
| **Sokoto** | 1342 (0.3%) | 3039 (0.6%) |  |
| **Taraba** | 4655 (1.1%) | 10702 (2.1%) |  |
| **Yobe** | 1912 (0.5%) | 3125 (0.6%) |  |
| **Zamfara** | 1218 (0.3%) | 1972 (0.4%) |  |

**Table 5:** Adjusted odds ratios for month of interruption and state where interruption occurred.

|  | **Adjusted odds ratios**<br>**(aOR)** | **95% Confidence Intervals**<br>**LCB** | **p-value** |
| --- | --- | --- | --- |
| **Month of interruption**<br>**(Ref=January)** |  |  |  |
| **February** | 0.44 | 0.43 - 0.46 | <.001 |
| **March** | 0.38 | 0.36 - 0.39 | <.001 |
| **April** | 0.25 | 0.24 - 0.26 | <.001 |
| **May** | 0.17 | 0.16 - 0.18 | <.001 |
| **June** | 0.21 | 0.20 - 0.22 | <.001 |
| **July** | 0.23 | 0.22 - 0.23 | <.001 |
| **August** | 0.62 | 0.60 - 0.65 | <.001 |
| **September** | 1.03 | 0.98 - 1.07 | 0.226 |
| **October** | 0.78 | 0.74 - 0.81 | <.001 |
| **November** | 1.16 | 1.11 - 1.21 | <.001 |
| **December** | 1.35 | 1.29 - 1.41 | <.001 |
| **State (Ref=Akwa Ibom)** |  |  |  |
| **Abia** | 1.36 | 1.24 - 1.49 | <.001 |
| **Adamawa** | 1.36 | 1.27 - 1.45 | <.001 |
| **Anambra** | 4.1 | 3.91 - 4.30 | <.001 |
| **Bauchi** | 3.5 | 3.29 - 3.73 | <.001 |
| **Bayelsa** | 1.98 | 1.86 - 2.10 | <.001 |
| **Benue** | 1.43 | 1.38 - 1.47 | <.001 |
| **Borno** | 1.5 | 1.40 - 1.60 | <.001 |
| **Cross River** | 1.95 | 1.87 - 2.02 | <.001 |
| **Delta** | 1.01 | 0.97 - 1.05 | 0.664 |
| **Ebonyi** | 1.41 | 1.26 - 1.58 | <.001 |
| **Edo** | 3.35 | 3.17 - 3.54 | <.001 |
| **Ekiti** | 1.19 | 0.99 - 1.42 | 0.064 |
| **Enugu** | 2.36 | 2.24 - 2.49 | <.001 |
| **FCT** | 1.7 | 1.63 - 1.77 | <.001 |
| **Gombe** | 1.51 | 1.42 - 1.61 | <.001 |
| **Imo** | 3.28 | 3.11 - 3.47 | <.001 |
| **Jigawa** | 2.33 | 2.12 - 2.56 | <.001 |
| **Kaduna** | 1.25 | 1.18 - 1.33 | <.001 |
| **Kano** | 2.98 | 2.82 - 3.13 | <.001 |
| **Katsina** | 1.22 | 1.12 - 1.32 | <.001 |
| **Kebbi** | 2.99 | 2.79 - 3.21 | <.001 |
| **Kogi** | 1.5 | 1.40 - 1.60 | <.001 |
| **Kwara** | 2.97 | 2.73 - 3.23 | <.001 |
| **Lagos** | 2.09 | 2.01 - 2.17 | <.001 |
| **Nasarawa** | 1.48 | 1.42 - 1.54 | <.001 |
| **Niger** | 2.67 | 2.53 - 2.82 | <.001 |
| **Ogun** | 2.75 | 2.59 - 2.92 | <.001 |
| **Ondo** | 1.02 | 0.93 - 1.13 | 0.678 |
| **Osun** | 1.96 | 1.76 - 2.19 | <.001 |
| **Oyo** | 2.49 | 2.32 - 2.67 | <.001 |
| **Plateau** | 1.29 | 1.20 - 1.39 | <.001 |
| **Rivers** | 2.17 | 2.12 - 2.23 | <.001 |
| **Sokoto** | 3.05 | 2.78 - 3.33 | <.001 |
| **Taraba** | 1.9 | 1.81 - 1.99 | <.001 |
| **Yobe** | 1.6 | 1.47 - 1.74 | <.001 |
| **Zamfara** | 1.88 | 1.7 - 2.08 | <.001 |

Logistic regression model: IIT = LW + VL + CD4 + Ambulatory + Bedridden + Unknown\_Functional\_Status + Male + No\_Education + Tertiary\_Education + Primary\_Education + Qranic\_Education + Unknown\_Education + Retired + Student + Unemployed + Unknown\_Employment + Other\_Marital\_Status + Single + Unknown\_Marital\_Status + Year\_Enrolled\_in\_Care + Surge + COVID + February + March + April + May + June + July + August + September + October + November + December + Urban + Abia + Adamawa + Anambra + Bauchi + Bayelsa + Benue + Borno + Cross\_River + Delta + Ebonyi + Edo + Ekiti + Enugu + Federal\_Capital\_Territory + Gombe + Imo + Jigawa + Kaduna + Kano + Katsina + Kebbi + Kogi + Kwara + Lagos + Nasarawa + Niger + Ogun + Ondo + Osun + Oyo + Plateau + Rivers + Sokoto + Taraba + Yobe + Zamfara + Age18\_24 + Age35\_44 + Age45\_54 + Age55\_64 + Age65ge + NNRTI + Undocumented\_ART + PI + WHO\_missing + WHO\_II + WHO\_III + WHO\_IV

**Table 6:** Adjusted hazard ratios (aHR) for month of interruption and state (location.

|  | **Adjusted hazard ratios (aHR)** | **95% confidence intervals** | **p.value** |
| --- | --- | --- | --- |
| **Month of interruption (Reference = January)** |  |  |  |
| **February** | 0.9 | 0.88 - 0.91 | <.001 |
| **March** | 0.79 | 0.78 - 0.80 | <.001 |
| **April** | 0.86 | 0.85 - 0.88 | <.001 |
| **May** | 0.7 | 0.69 - 0.72 | <.001 |
| **June** | 0.76 | 0.75 - 0.77 | <.001 |
| **July** | 0.83 | 0.82 - 0.85 | <.001 |
| **August** | 1.18 | 1.16 - 1.19 | <.001 |
| **September** | 1.2 | 1.18 - 1.22 | <.001 |
| **October** | 1.11 | 1.09 - 1.12 | <.001 |
| **November** | 1.22 | 1.20 - 1.24 | <.001 |
| **December** | 1.19 | 1.17 - 1.21 | <.001 |
| **State (Reference=Akwa Ibom)** |  |  |  |
| **Abia** | 0.74 | 0.71 - 0.78 | <.001 |
| **Adamawa** | 0.95 | 0.92 - 0.98 | 0.001 |
| **Anambra** | 0.98 | 0.96 – 1.00 | 0.106 |
| **Bauchi** | 1.52 | 1.48 - 1.55 | <.001 |
| **Bayelsa** | 1.03 | 1.00 - 1.07 | 0.075 |
| **Benue** | 0.85 | 0.83 - 0.86 | <.001 |
| **Borno** | 1.11 | 1.07 - 1.14 | <.001 |
| **Cross River** | 1.09 | 1.07 - 1.11 | <.001 |
| **Delta** | 0.78 | 0.76 - 0.80 | <.001 |
| **Ebonyi** | 1.26 | 1.18 - 1.35 | <.001 |
| **Edo** | 1.37 | 1.34 - 1.40 | <.001 |
| **Ekiti** | 0.81 | 0.73 - 0.90 | <.001 |
| **Enugu** | 1.25 | 1.21 - 1.30 | <.001 |
| **FCT** | 0.81 | 0.79 - 0.82 | <.001 |
| **Gombe** | 0.87 | 0.84 - 0.90 | <.001 |
| **Imo** | 1.37 | 1.33 - 1.41 | <.001 |
| **Jigawa** | 1.35 | 1.30 - 1.40 | <.001 |
| **Kaduna** | 1.21 | 1.17 - 1.25 | <.001 |
| **Kano** | 1.16 | 1.14 - 1.19 | <.001 |
| **Katsina** | 0.71 | 0.68 - 0.74 | <.001 |
| **Kebbi** | 0.58 | 0.56 - 0.59 | <.001 |
| **Kogi** | 1.05 | 1.02 - 1.08 | 0.002 |
| **Kwara** | 0.79 | 0.76 - 0.82 | <.001 |
| **Lagos** | 0.96 | 0.94 - 0.98 | <.001 |
| **Nasarawa** | 0.97 | 0.95 - 0.99 | 0.015 |
| **Niger** | 0.81 | 0.79 - 0.83 | <.001 |
| **Ogun** | 1.07 | 1.04 - 1.10 | <.001 |
| **Ondo** | 0.77 | 0.73 - 0.82 | <.001 |
| **Osun** | 0.93 | 0.87 - 0.98 | 0.014 |
| **Oyo** | 1.36 | 1.32 - 1.40 | <.001 |
| **Plateau** | 1.23 | 1.20 - 1.27 | <.001 |
| **Rivers** | 1.02 | 1.01 - 1.04 | 0.008 |
| **Sokoto** | 0.99 | 0.95 - 1.02 | 0.491 |
| **Taraba** | 0.55 | 0.54 - 0.56 | <.001 |
| **Yobe** | 1.13 | 1.09 - 1.18 | <.001 |
| **Zamfara** | 0.68 | 0.65 - 0.71 | <.001 |

Cox proportional hazards model: Time to IIT event = Last\_Redorded\_Weight + Viral\_Load\_Recorded + CD4\_Recorded + Ambulatory + Bedridden + Unknown\_Fucntional\_Status + Male + No\_Education + Tertiary + Primary + Qranic\_Education + Unknown\_Education + Retired + Student + Unemployed + Unknown\_Employment + Other\_Marital\_Status + Single + Unknown\_Marital\_Status + Year\_enrolled\_in\_Care + Surge + COVID + February + March + April + May + June + July + August + September + October + November + December + Urban + Abia + Adamawa + Anambra + Bauchi + Bayelsa + Benue + Borno + Cross\_River + Delta + Ebonyi + Edo + Ekiti + Enugu + Federal\_Capital\_Territory + Gombe + Imo + Jigawa + Kaduna + Kano + Katsina + Kebbi + Kogi + Kwara + Lagos + Nasarawa + Niger + Ogun + Ondo + Osun + Oyo + Plateau + Rivers + Sokoto + Taraba + Yobe + Zamfara + Age18\_24 + Age35\_44 + Age45\_54 + Age55\_64 + Aage65ge + NNRTI + Undocumented\_ART + PI + WHO\_missing + WHO\_II + WHO\_III + WHO\_IV

